# Suitability of Deep Weakly Supervised Learning to detect Acute Ischemic Stroke and Hemorrhagic Infarction Lesions Using Diffusion-weighted Imaging

**DOI:** 10.1101/2020.03.05.20031484

**Authors:** Chen Cao, Zhiyang Liu, Guohua Liu, Song Jin, Shuang Xia

## Abstract

**Objectives:** The automatic detection of acute ischemic stroke (AIS) and hemorrhagic infarction (HI) based on deep learning could avoid missed diagnosis. The fully supervised learning requires the amount of time and the expertise to manually outline lesions, which limits its applicability. The weakly supervised learning has the potential to reduce the labeling workload. The purpose of this study was to evaluate a weakly supervised method in detection of AIS and HI location using DWI.

**Methods:** We proposed to adopt weakly supervised learning to spatially-locate AIS lesions by residual neural network (ResNet) and visual geometry group (VGG) network. On an AIS dataset, the area under the receiver operating characteristic (ROC) curve (AUC), sensitivity and precision were calculated. Next, ResNet, which presented superior performance on the AIS dataset, was further applied to an HI dataset.

**Results:** In the AIS dataset, the AUCs of ResNet and VGG on identifying image slices with AIS were 0.97 and 0.94, respectively. On spatially-locating the AIS lesions, ResNet provided higher sensitivity and a lower missed diagnosis rate than VGG, especially for pontine AIS lesions. In the HI dataset, the sensitivity of ResNet was 87.73% for AIS detection, and 86.20% for HI detection, respectively.

**Conclusions:** Weakly supervised learning can effectively detect the location of AIS and HI lesions in DWI, which is of paramount importance in avoiding misdiagnosis in clinical scenario.

**Key points:** The deep weakly supervised learning can reduce the labeling workload; ResNet can obtain more exact results, especially for pontine AIS lesions; Weakly supervised learning can effectively detect AIS and HI lesions in DWI

## Introduction

Acute ischemic stroke (AIS) rank as the first leading cause of death in China. The age standardized prevalence, incidence, and mortality rates of AIS in China are 111,4.8, 246.8 and 114.8/100,000 person-years.[1] The latest guidelines recommend imaging evaluations before and after treatment, with particular attention being given to the occurrence of hemorrhagic transformation (HT).[2,3] Studies have shown that the tendency of HT in Asian population is significantly higher than that in Western population.[4] Whether spontaneous HT or therapy-related HT, missed diagnosis should be avoided to affect the prognosis of patients.[5] However, in China, the uneven development of medical level in different regions leads to the uneven imaging diagnosis ability of clinicians. If there is a professional method to accurately mark the location of AIS and HT lesions, it will help clinicians avoid missed diagnosis.

According to the most widely used classification of HT from the European Cooperative Acute Stroke Study II (ECASS II), HT is classified into two broad categories: hemorrhagic infarction (HI) and parenchymal hematoma (PH).[6] An HI indicates the presence of petechiae within the infarcted area without a space-occupying effect, which is more difficult to find than PH. Although HI is usually benign, some studies have found that an HI is actually a negative predictor of a good outcome.[7,8]

Diffusion weighted imaging (DWI), including diffusion images, apparent diffusion coefficient (ADC) and b_0_ images, should be routinely obtained for patients who are clinically suspected of having AIS lesions. Although b0 images are more difficult to see with the naked eye than gradient recalled echo (GRE) when detecting HI lesions, obtaining the GRE normally takes several minutes, which can prolong the examination time for these AIS patients.[9-11] Some studies has found no significant difference in the sensitivity between b0 images and GRE for acute hemorrhage detection.[12,13]

Many fully supervised methods, such as EDD-Net[14], 3D-DenseNet[15] and the residual-structured fully convolutional network (Res-FCN)[16], have been successfully applied in AIS lesion detection. However, the fully supervised approaches require the accurate annotation of lesion outlines. The accurate annotation of lesions for subjects requires a substantial amount of time, which limits its applicability. Recently, some weakly supervised methods were proposed to leverage the annotation workload[17], such as the wiseDNN[18] and the 3D weakly supervised CNN[19]. The above articles have indicated that weakly supervised methods can reduce the difficulty of label acquisition while still maintaining high detection efficiency.

We hypothesized that weakly supervised learning can sensitively detect the location of AIS and HT lesions based on DWI. Therefore, the purpose of the current paper is to evaluate a weakly supervised method in detection of AIS and HI location using DWI.

## Methods

### Experiment on acute ischemic stroke data

Ethical approval was granted by the Tianjin Huanhu Hospital Medical Ethics Committee. All clinical images were collected from a retrospective database and anonymized prior to use. In this study, the MR images of 736 subjects with AIS were collected from Tianjin Huanhu Hospital. The eligibility for inclusion in this study was AIS within 3 days from the onset of symptoms and available diffusion-weighted images, between January 1, 2017, and October 30, 2018. MRI acquisition were provided in the Data Supplement.

The whole dataset was divided into training set and test set. The training set included all 417 weakly annotated subjects, and was used for training the neural network and fine-tune the hyper-parameters. The subjects in the training dataset were randomly sampled from the retrospective database. The test set included 319 fully-annotated subjects, and was used only for evaluating the performance. To detect whether weakly supervised learning is affected by the size and location of the lesion and evaluate the performance of the weakly supervised learning on the small lesions, the subjects in the test set were carefully selected. In particular, 240 subjects had lacunar infarction (LI) and 79 had territorial infarction (TI). A subject was categorized as an LI subject if the lesion was singular and its volume was smaller than 1766 mm^3^, i.e., the volume of a ball with a radius of 7.5 mm. Among the 240 LI subjects, we further divided them into 3 groups according to the lesion positions on the basal ganglia (LI-B), pons (LI-P) and centrum semiovale (LI-C), with 80 subjects in each group.

Fig. 1A showed the differences between weak labels and full labels. The labels were annotated by an experienced expert (Dr. Chen Cao, a radiologist with 8 years of neuroimaging experience) in the DWI b=1000 image, while also referring to the ADC. The expert (Dr. Chen Cao) performed the manual annotation of the train and test data twice. If the interrater reliability was high, it would prove its credibility. Manual annotated of the first trial served as the ground-truth. Another experienced expert checked the labels (Dr. Song Jin, a radiologist with 20 years of neuroimaging experience). More details of the patient characteristics and consistency test were provided in the Data Supplement.

**Fig. 1.**
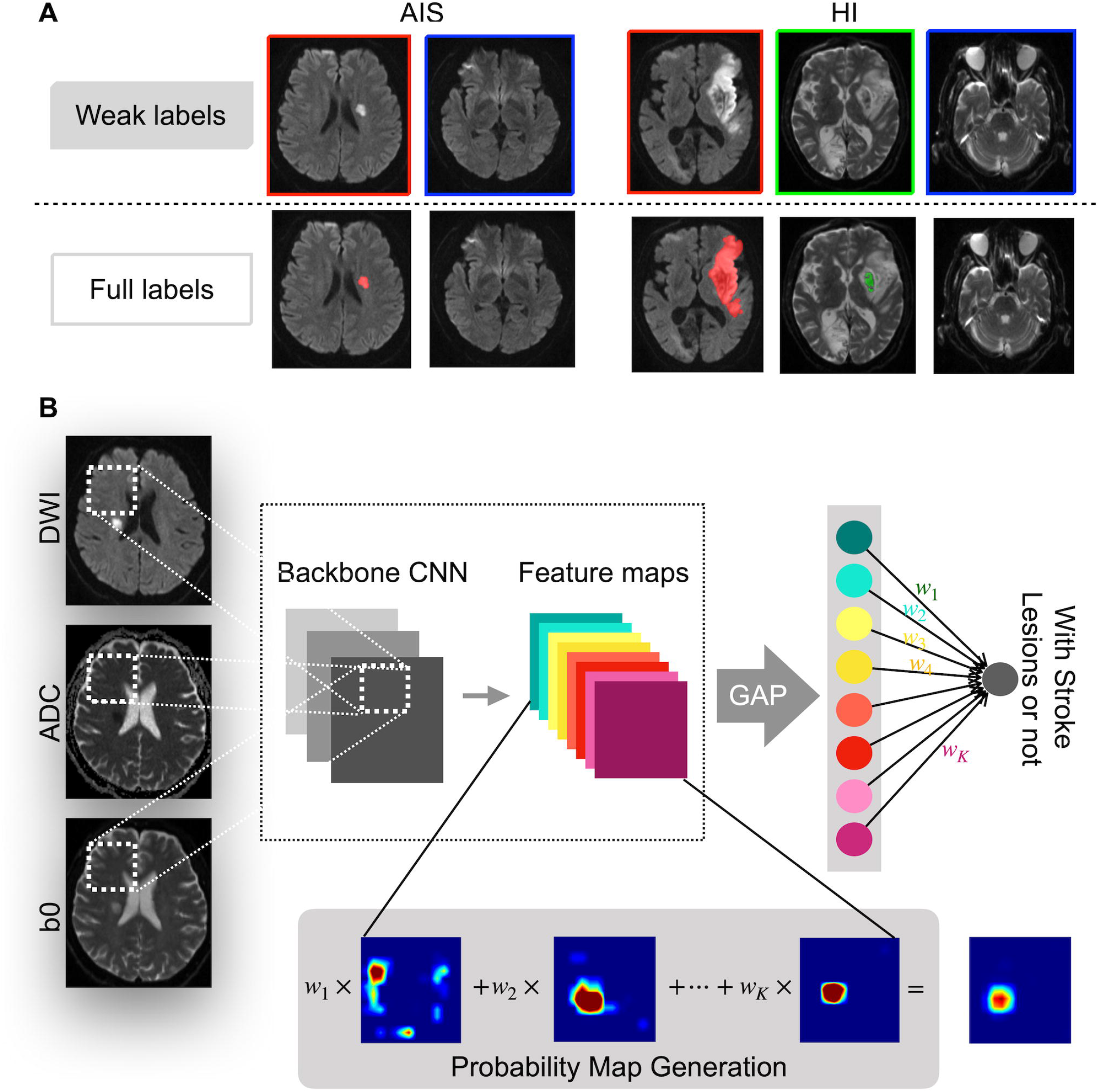
A. A task of classifying healthy (blue) vs acute ischemic stroke (AIS) (red) and hemorrhagic infarction (HI) (green) tissue in DWI images. With the weak labels, each slice of the subjects was only annotated as “with lesion” or “without lesion”. With full labels, however, all the stroke lesions were annotated in a pixel-by-pixel manner. B. Network architecture for lesion detection by using the weakly supervised learning method. CNN, convolutional neural network; GAP, global average pooling

### Experiment on hemorrhage infarction data

We further collected 305 AIS subjects with HI lesions from the retrospective database. Two hundred and forty subjects with weak annotations were randomly included in the training set, and 65 subjects with full annotations were in the test set. Each image slice in the training set included two annotations: with or without an AIS lesion and with or without an HI lesion. The HI lesions were manually annotated by inspecting the GRE images. The Data Supplement provides the details of the patient characteristics.

In our experiment, we used DWI, ADC and b0 images as input. The results of the HI detection by using GRE were supposed to be the gold standard. The process of labeling lesions is similar to that of the AIS data. To simultaneously detect both HI and AIS lesions, we modified the network in Fig. 1B by using a fully connected (FC) layer with 2 neurons as the output layer. The network was then trained by following the approach that was described in the network setup.

### CNN Architecture

In our experiments, we choose to use the VGG and ResNet networks as the convolutional neural network (CNN) backbone. A CNN was used to extract the features. Conventionally, a CNN was designed for classification, and therefore the last convolution layer was followed by a fully connected layer to output the classification result, as shown in Fig. 1B.In the training stage, as the manual annotations denoted whether a slice contains a stroke lesion or not, we used a CNN classifier to train the network. The feature maps in the last convolution layer were processed by a global average pooling (GAP) layer, which outputs the mean value of each feature map. The mean values were further processed by a dense layer for classification. In the testing stage, as our objective was to localize the stroke lesions, we needed to convert the trained CNN classifier such that the network can output an attention map that shows where the lesions were located. We directly output the feature maps of the last convolutional layer, and used the weighted sum as the localization results to generate a class activation map (CAM).

To evaluate the performance of the CAM-based methods, we proposed several lesion-wise metrics using 3D connected component analysis. We measured the per-subject mean numbers of false positive lesions (mFP-L), false negative lesions (mFN-L). Next, we further defined the lesion-wise sensitivity (i.e. recall) and precision (i.e. positive predictive value) to evaluate the lesion-wise performance. In addition, the subject-wise detection rate also matters in clinical diagnosis. We used the number of failed detected subjects (FD-S) to evaluate the subject-level performance. More details of the CNN architecture and evaluation metrics were provided in the Data Supplement.

### Implementation

The experiments were performed on a computer with an Intel Core i7-7800K CPU, 64GB RAM and Nvidia GeForce 1080Ti GPU with 11GB memory. The computer operated on Ubuntu Linux 16.04 LTS with CUDA 9.0. The network was implemented on Keras 2.0.2. The MR image files were stored as Neuroimaging Informatics Technology Initiative (NIfTI) format, and processed using Simple Insight ToolKit (SimpleITK) [3.8.0]. We used ITK-SNAP [3.8.0] for image annotation and the visualization of results.

### Statistical Analysis

Statistical analyses were performed using MedCalc for Windows software version 19.0.2 (MedCalc software, Mariakerke, Belgium). To verify the consistency of the labels that were given by experts twice, the intraclass correlation coefficient (ICC) and *k* coefficient was computed between the 2-lesion measurements. The strength of agreement of the ICC and *k* coefficient values was categorized as follows: poor (<0.20), fair (0.21∼0.40), moderate (0.41∼0.60), good (0.61∼0.80), and excellent (>0.80). The receiver operating characteristic curves (ROCs) and the area under the curves (AUCs) were calculated to compare the efficacy of each model. First, a 2-paired-sample Wilcoxon test was performed to determine if the VGG and ResNet were significantly different in terms of parameters such as the sensitivity, precision, mFP-L and mFN-L. A Kruskal-Wallis test and multiple comparisons were performed to determine if the TIs, LI-Bs, LI-Ps and LI-Cs of the VGG and ResNet were significantly different. Then, a 2-independent-sample Mann-Whitney test was performed to determine the difference in AIS and HI lesions that were detected by the previous best method. *P* values <0.05 were considered to indicate statistical significance.

## Results

### Acute ischemic stroke data

We first evaluated the network classification performance by evaluating the ROC curves, as shown in Fig. 2A. It was clear from Fig. 2A that both classifiers possessed high performance for identifying image slices with AIS lesions. In particular, the ResNet achieved an AUC score of 0.974, which highlighted the great potential of this deep learning method to aid clinicians’ diagnoses.

**Fig. 2.**
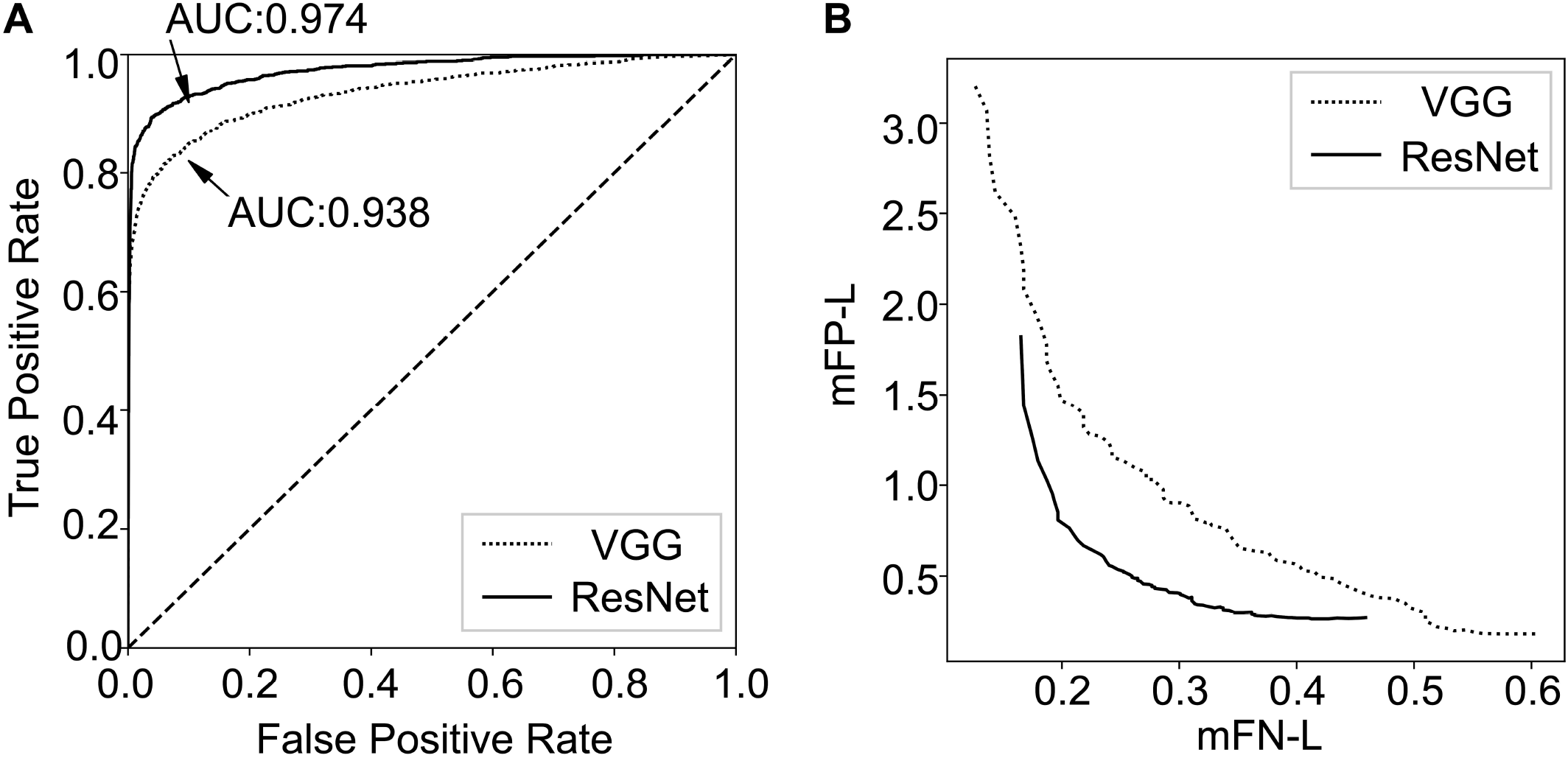
A. The receiver operating characteristic (ROC) curve of the residual neural (ResNet) and visual geometry group (VGG) network classifiers shows the false positive rate (x-axis) vs. the true positive rate (y-axis). The areas under the ROC curve (AUCs) for the ResNet and VGG networks were both superior in being able to identify lesions in acute ischemic stroke (AIS) image slices. B. The performance of the weakly supervised lesion detection method in being able to detect lesions shows the dependence between the mean number of false-negative lesions (mFN-L) (x-axis) vs. false-positive lesions (mFP-L) (y-axis) in the AIS test dataset

Note that the lesion-wise performance was highly related to the choice of the threshold. Intuitively, as the threshold increases, the lesions on the predicted segmentation maps shrink, leading to more FN-Ls and a smaller amount of FP-Ls. Fig. 2B plots the dependence between mFN-Ls and mFP-Ls of ResNet and VGG on the test dataset. As the output for the pixel *x*_*i*_, ∈ [0, 1] which can be interpreted as the probability that the pixel is classified as lesion tissue, a threshold δ is used to generate the final binary segmentation, where the final binary output *x*_*i*_ = 1 if *x*_*i*_ > δ, and *x*_*i*_ = 0 otherwise. Each dot of the curve was plotted by using different values of the threshold δ, ranging from 0.5 to 1. The tradeoffs of ResNet and VGG were also plotted for comparison. As the threshold δ increases, the mFP-L decreased at the expense of a higher mFN-L. As we can see, the ResNet performed better with respect to the tradeoff curves.

In our work, we used the slice-level label to perform lesion-level localization and detection. Fig. 3 presents some examples of the class activation map of AIS lesions. The original DWI, the ADC map and the manual segmentation were also presented for comparison. As we could observe from Fig. 3, both VGG and ResNet were able to highlight the lesions. The ResNet, however, tended to be more precise in outlining the lesions.

**Fig. 3.**
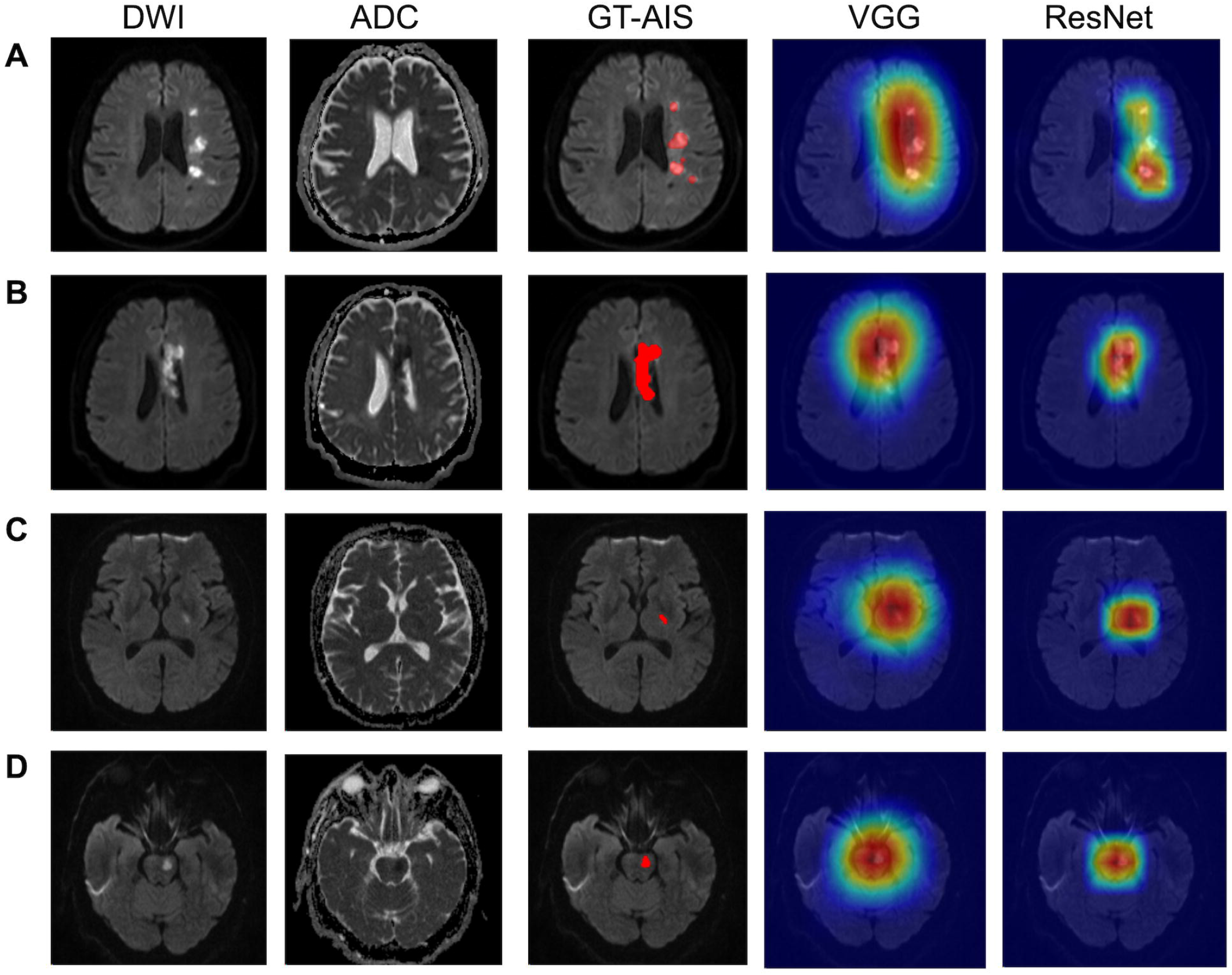
Lesion detection results using the weakly supervised method on 319 patients with acute ischemic stroke (AIS) lesions. Four patient examples are shown (A, B, C, D). The AIS lesions were annotated in red in the ground truth (GT) images. Subjects A and B belonged to the territorial infarction groups, with multiple or large lesions. Subjects C and D belonged to the lacunar infarction groups, which had lesions that were located in the basal ganglia and pons, respectively. DWI, diffusion-weighted imaging; ADC, apparent diffusion coefficient; and ResNet, residual neural network; and VGG, visual geometry group network

Note that the output of the CNN indicated the probability that a pixel should be labeled as lesion tissue, and a threshold δ was required to convert the probability score map to a binary segmentation. The threshold was selected by maximizing the F1 score on the validation set. Table 1 summarizes the numerical evaluation results. We evaluated the performance for 4 groups of subjects: the TI group, the LI-B group, the LI-P group, and the LI-C group. Table 1 shows that ResNet effectively reduced the mFN-L, mFP-L and FD-S compared with VGG. Notably, for the LI-P group, FD-S decreased from 14 to 1. For the LI-B group, the small difference in the mFN-Ls of the two methods was due to its small sample size, which made it impossible to estimate the *P* values. For both methods, the mFN-L for the LI-C group was 0. Table 1 also shows that the sensitivity of the TI group was lower than that of the LI groups (*P*<0.05), regardless of whether ResNet or VGG was used. In VGG, the sensitivity of the LI-P group was lower than that of the LI-B and LI-C groups (*P*<0.05). ResNet improved the detection performance of the LI-P group, making sensitivity of the LI groups more than 95%. In VGG, the precisions of the LI-P and LI-C groups were lower than that of the LI-B group (*P*<0.05). In ResNet, there is no significant difference in the results of LI detection among the three groups.

**Table 1.** Lesion-wise performance of VGG and ResNet on the acute ischemic stroke (AIS) test sets

|  | TI |  |  | LI-B |  |  | LI-P |  |  | LI-C |  |  |
| --- | --- | --- | --- | --- | --- | --- | --- | --- | --- | --- | --- | --- |
| Method | VGG | ResNet | *P* | VGG | ResNet | *P* | VGG | ResNet | *P* | VGG | ResNet | *P* |
| mFN-L | 1.63 | 1.29 | 0.11 | 0.05 | 0.04 | — | 0.21 | 0.04 | 0.00 | 0 | 0 | — |
| mFP-L | 0.81 | 0.30 | 0.00 | 0.56 | 0.25 | 0.01 | 0.81 | 0.13 | 0.00 | 0.85 | 0.18 | 0.00 |
| FD-S | 2 | 0 |  | 0 | 0 |  | 14 | 1 |  | 0 | 0 |  |
| Sensitivity | 68.06% | 74.75% | 0.13 | 95.36% | 96.47% | — | 79.52% | 96.39% | 0.00 | 100% | 100% | — |
| Precision | 81.12% | 92.63% | 0.00 | 64.29% | 80.39% | 0.03 | 50.38% | 88.89% | 0.00 | 54.05% | 85.11% | 0.00 |

mFN-L, mean number of false negative lesions; mFP-L, mean number of false positive lesions; FD-S, the number of failure-to-detect subjects; TI, territorial infarction group, LI-P lacunar-lesion (pons) group; LI-B, basal ganglia group; LI-C, semi oval center group; ResNet, residual neural network; and VGG, visual geometry group network.

### Hemorrhage Infarction data

Therefore, due to ResNet’s excellent performance in AIS detection, it was further applied to HI detection. Table 2 summarizes the numerical evaluation results of HI lesions. The results show that the indicators of each detection parameter reached a high level, which is consistent with the detection effectiveness of AIS lesions and has no significant difference. However, one case of AIS in the pons was missed.

**Table 2.** Lesion-wise performance of ResNet on the hemorrhagic infarction (HI) test sets

|  | AIS | HI | *P* |
| --- | --- | --- | --- |
| mFN-L | 0.78 | 0.68 | 0.35 |
| mFP-L | 0.37 | 0.43 | 0.53 |
| FD-S | 1 | 1 |  |
| Sensitivity | 87.73% | 86.20% | 0.67 |
| Precision | 91.38% | 87.61% | 0.38 |

AIS, acute ischemic stroke; mFN-L, mean number of false negative lesions; mFP-L, mean number of false positive lesions; FD-S, the number of failure-to-detect subjects

Fig. 4 shows that the trained ResNet can annotate the AIS and HI lesions in the data. It is difficult for us to determine the existence of HI lesions by using naked eye observations of b0 images, and this finding needs to be confirmed by GRE. However, weakly supervised learning accurately detected the location of AIS and HI lesions in DWI. Subject C had an old hemorrhagic lesion in the contralateral hemisphere of the AIS lesion, which was not misdiagnosed as an HI.

**Fig. 4.**
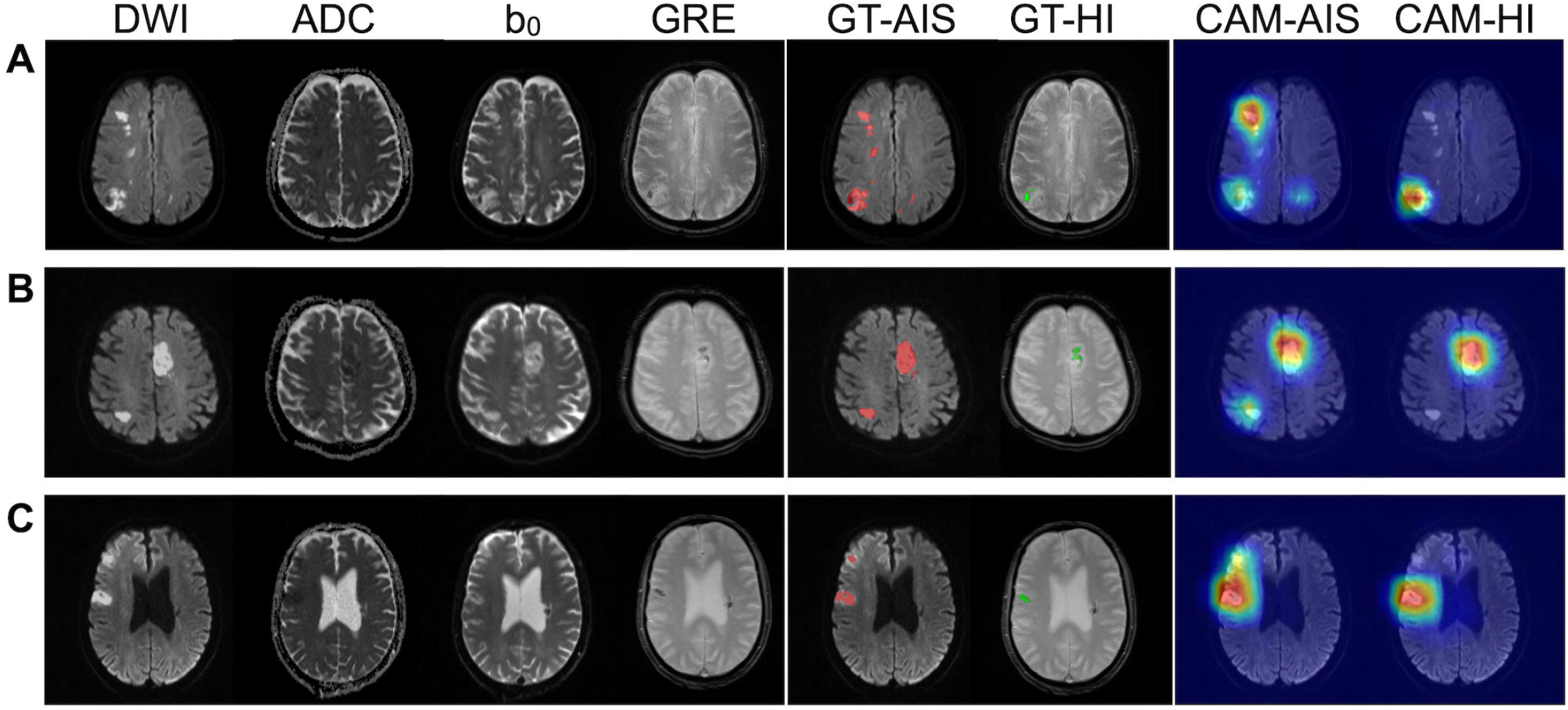
Three examples of hemorrhagic infarction (HI) patient imaging (A, B, and C). In the ground truth (GT) images, the acute ischemic strokes (AIS) lesions and the HI lesions were annotated in red and green, respectively. CAM, class activation map

## Discussion

The contributions of this paper were summarized as follows. First, a weakly supervised method was proposed. ResNet improved the sensitivity of stroke lesion detection and reduced the mFN-L and mFP-L, especially for pontine lesions, which highlighted the importance of utilizing a residual structure. Second, the proposed method can automatically and sensitively detect AIS and HI lesions based on DWI.

In recent years, the application of deep learning to lesion detection in medical imaging has become a popular topic due to its high processing efficiency and analysis speed.[20] Some studies have accurately detected AIS lesions using fully supervised methods. The accurate annotation of AIS lesions from a large number of images requires tremendous time.[21] In this paper, we proposed a weakly supervised method to detect lesions, which was very effective for detecting AIS lesions. ResNet had more advantages than VGG. It was worth noting that the FD-S of the LI-P group in VGG was actually as high as 14. This result may be due to the influence of magnetically susceptible artifacts that were caused by the inhomogeneous magnetic field due to the magnetic susceptibility differences between the brain tissue and air-containing areas of the skull.[22] ResNet improved the sensitivity and precision of the LI-P group and reduced the FD-S to 1. In addition, the precision of the LI-C group in VGG was lower than that of the LI-B group. This result may be due to the frequent occurrence of T_2_ shine-through in the centrum semiovale that is caused by the demyelination of white matter.[23] ResNet also solved this problem well, increasing the precision from 50.05% to 85.11%. ResNet can make the network much deeper and avoid the vanishing gradient problem.[24] Instead of simply stacking more convolution layers, many skip connections are added between the layers. Such a structure allows the gradient to pass backward through the skip connections, and all the convolution layers are able to be updated to extract features after the first training epoch. Consequently, ResNet can extract deeper image features and obtain more exact results. Table 2 suggested that the sensitivity in the TI test sample was low. Because the lesions in LI groups were single and regular in shape, while the lesions in TI group were often multiple and irregular. Small dotted satellite foci appeared around large lesions, so mFN-L increased naturally. However, FD-S was 0, indicating no missed diagnosis. Therefore, this result will not limit its clinical utility.

HI may occur as part of the spontaneous evolution of AIS, or they be precipitated by the use of antiplatelet, anticoagulant, or thrombolytic therapy.[25] A spontaneous HI can occur prior to any treatment and may increase the amount of bleeding and seriously influence the patient prognosis if it is not detected and treated with thrombolysis in a timely manner.[26] Therefore, identifying spontaneous HI during pretreatment stages is helpful for guiding the selection of patients for whom thrombolytic or antithrombotic treatments would be appropriate. However, therapy-related HI, i.e., posttherapy HI, is a medical emergency, with deterioration typically occurring quickly after onset. ResNet was trained on an AIS dataset, and it achieved high detection accuracy. The method was also applied to an HI dataset, and the sensitivities were 87.73% and 86.20% for AIS and HI lesions, respectively. Therefore, our proposed method was sensitive to both AIS and HI lesions.

There are several limitations in the present study. First, the lesion segmentation accuracy needs to be further improved. In this paper, due to the data training of the weakly supervised learning, the information that is provided by weakly supervised labels is significantly less than that provided by fully supervised labels. Although this method can accurately mark the locations of lesions, it is difficult to provide detailed parameters such as the size of a lesion. Second, although some examples can prove that this method can distinguish HI lesions from hemosiderin deposits, it has not been systematically confirmed and needs further verification. This method still depends on the clinical history and the changes in the patients’ symptoms. Third, note that the lesion-wise specificity was not included in our analysis, due to the fact that the specificity is a true-negative-related metric, and it is hard to define a true-negative lesion in our task. Fourth, in order to further evaluate the detection accuracy of the proposed method, multicenter studies are warranted that also standardize imaging protocols as well as post-processing procedures.

## Conclusions

The weakly supervised learning could automatically and sensitively detect location of AIS and HI lesions based on DWI, thus effectively avoiding missed diagnosis. The proposed method helps to reduce the problem of insufficient medical imaging data and the difficulty of obtaining expert labels.

## Data Availability

Some or all data, models, or code generated or used during the study are available from the corresponding author by request.

## Abbreviations and acronyms

ResNet: residual neural network
VGG: visual geometry group
mFP-L: mean number of false positive lesions
mFN-L: mean number of false negative lesions
FD-S: failed detected subjects
LI: lacunar infarction
TI: territorial infarction

